# Regional 4D Cardiac Magnetic Resonance Strain Predicts Cardiomyopathy Progression in Duchenne Muscular Dystrophy

**DOI:** 10.1101/2023.11.07.23298238

**Authors:** Conner C. Earl, Alexa M. Jauregui, Guang Lin, Kan N. Hor, Larry W. Markham, Jonathan H. Soslow, Craig J. Goergen

**Affiliations:** Weldon School of Biomedical Engineering, Purdue University, West Lafayette, IN; Indiana University School of Medicine, Indianapolis, IN; Department of Mathematics & School of Mechanical Engineering, Purdue University, West Lafayette, IN; The Heart Center, Nationwide Children’s Hospital, Ohio State University, Columbus, OH, USA; Division of Pediatric Cardiology, Riley Children’s Hospital at Indiana University Health, Indiana University School of Medicine, Indianapolis, IN; Division of Pediatric Cardiology, Department of Pediatrics, Vanderbilt University Medical Center

**Keywords:** Duchenne muscular dystrophy, cardiomyopathy, strain, gadolinium, progression, Left ventricular function, Diastolic dysfunction

## Abstract

**Background:** Cardiomyopathy (CMP) is the leading cause of death in Duchenne muscular dystrophy (DMD). Characterization of disease trajectory can be challenging, especially in the early stage of CMP where onset and clinical progression may vary. Traditional metrics from cardiovascular magnetic resonance (CMR) imaging such as LVEF (left ventricular ejection fraction) and LGE (late gadolinium enhancement) are often insufficient for assessing disease trajectory. <u>We hypothesized that strain patterns</u> from a novel 4D (3D+time) CMR regional strain analysis method <u>can be used to predict</u> <u>the rate of DMD CMP progression.</u>

**Methods:** We compiled 115 short-axis cine CMR image stacks for n=40 pediatric DMD patients (13.6±4.2 years) imaged yearly for 3 consecutive visits and computed regional strain metrics using custom-built feature tracking software. We measured regional strain parameters by determining the relative change in the localized 4D endocardial surface mesh using end diastole as the initial reference frame.

**Results:** We first separated patients into two cohorts based on their initial CMR: LVEF≥55% (n=28, normal cohort) and LVEF<55% (n=12, abnormal cohort). Using LVEF decrease measured two years following the initial scan, we further subclassified these cohorts into slow (ΔLVEF%≤5) or fast (ΔLVEF%>5) progression groups for both the normal cohort (n=12, slow; n=15, fast) and the abnormal cohort (n=8, slow; n=4, fast). There was no statistical difference between the slow and fast progression groups in standard biomarkers such as LVEF, age, or LGE status. However, basal circumferential strain (E_cc_) late diastolic strain rate and basal surface area strain (E_a_) late diastolic strain rate magnitude were significantly decreased in fast progressors in both normal and abnormal cohorts (*p*<0.01, *p*=0.04 and *p*<0.01, *p*=0.02, respectively). Peak E_a_ and E_cc_ magnitudes were also decreased in fast progressors, though these only reached statistical significance in the normal cohort (*p*<0.01, *p*=0.24 and *p*<0.01, *p*=0.18, respectively).

**Conclusion:** Regional strain metrics from 4D CMR can be used to differentiate between slow or fast CMP progression in a longitudinal DMD cohort. These results demonstrate that 4D CMR strain is useful for early identification of CMP progression in patients with DMD.

**Clinical Perspective:** Cardiomyopathy is the number one cause of death in Duchenne muscular dystrophy, but the onset and progression of the disease are variable and heterogeneous. In this study, we used a novel 4D cardiovascular magnetic resonance regional strain analysis method to evaluate 40 pediatric Duchenne patients over three consecutive annual visits. From our analysis, we found that peak systolic strain and late diastolic strain rate were early indicators of cardiomyopathy progression. This method offers promise for early detection and monitoring, potentially improving patient outcomes through timely intervention and management.

## Introduction

Duchenne muscular dystrophy (DMD) is a devastating X-linked genetic disease characterized by progressive muscle weakness, loss of mobility, and often a dramatically shortened lifespan^1–3^. Muscle weakness and loss of ambulation are hallmark features of this disease, however, DMD-associated cardiomyopathy (CMP) is the number one cause of death in this population^4, 5^. While there is no cure for DMD, advancements in research and medical care have enhanced the quality of life and extended the life expectancy of individuals affected by this condition^4, 6^. Among the various developments in the management of DMD, cardiac magnetic resonance imaging (CMR) has emerged as the gold standard tool for assessing cardiac involvement and tracking the progression of this disease^7–10^. However, given the inherent variability in the onset and clinical progression of DMD, characterizing the trajectory of the disease, particularly in the early stages of CMP, presents a notable challenge^11^.

CMR-derived metrics most often used in disease assessment include left ventricular ejection fraction (LVEF) and late gadolinium enhancement (LGE)^7^. LVEF is a well-accepted indicator of overall systolic function, but a decrease in LVEF below clinically normal levels often occurs only after substantial disease progression^11^. LGE is an indicator of early disease and correlates with physiologic fibrofatty replacement of heart muscle tissue in DMD^12^. However, LGE might also be considered a relatively late finding as it can only be detected when a substantial portion of the myocardium has been affected. Cardiac strain measures the relative deformation of the myocardium throughout the cardiac cycle with end diastole assumed to be the reference state. Strain has shown promise as an early indicator of disease,^8^ but is not currently used extensively in clinical decision making.

We have recently developed novel methods for assessing regional strain using stacked short-axis CMR images^13^. This innovative approach involves the utilization of a method for quantifying 4D (3D+time) cardiovascular magnetic resonance (CMR) regional strain. With this approach, we hypothesize DMD patient groups can be differentiated based on their strain patterns to predict the rate of CMP progression, contributing to a deeper understanding of the disease, paving the way for more targeted interventions, and eventually improving patient outcomes.

## Methods

### Patient Population

In this study, we analyzed data from patients with a range of DMD-associated CMP from an observational study approved by the Vanderbilt Institutional Review Board. All participants adhered to approved protocols and provided their consent or assent. The original study enrolled DMD CMP patients who had received a phenotypic diagnosis of DMD, subsequently confirmed through genetic testing or muscle biopsy. As possible, each patient in the study had three consecutive CMR scans each about 12 months apart (12.3±0.8 months; mean±SD). Our study excluded patients with diagnoses other than DMD and those with inadequate assessment or non-diagnostic results for LGE.

### Patient Stratification

DMD patients presented with a range of CMP severity at the initial visit: 12 with reduced LVEF of <55% (30% of total) and 26 with LGE (65% of total). We initially categorized patient CMP into Stage A (LVEF≥55%, no LGE; n=14), Stage B (LVEF≥55%, with LGE; n=14), Stage C (40%<LVEF<55%, with LGE; n=9), or Stage D (LVEF<40%, with LGE; n=3) based on the 2022 AHA’s guideline for the management of heart failure^14^. We also grouped patients based on initial visit LVEF and progression after two years. We chose to group patients this way in an effort to model the natural history differences between DMD patients with normal LVEF (≥55%) vs. those with abnormal LVEF (<55%). The patient groups were further categorized by decrease in LVEF two years following the initial visit as follows: normal, slow progression: initial LVEF≥55%, +2 years ΔLVEF%≤5 (n=12), normal, fast progression: initial LVEF≥55%, +2 years ΔLVEF%>5 (n=15), abnormal, slow progression: initial LVEF<55% +2 years ΔLVEF%≤5 (n=8), abnormal, fast progression: initial LVEF<55%, +2 years ΔLVEF%>5 (n=4). One patient was excluded due to poor image quality.

### Image Acquisition

We obtained CMR images using a 1.5 Tesla scanner (Avanto or Avanto Fit, Siemens Healthineers, Erlangen, Germany) as described previously^13, 15^. Briefly, we acquired balanced steady-state free precession (bSSFP) cine images in the 4-chamber, 3-chamber, 2-chamber, and a short-axis stack. Typical imaging parameters were 6-8 mm thickness, 20-25 images per cardiac cycle with 11-17 short axis cine slices per patient. LGE imaging included single-shot phase-sensitive inversion recovery bSSFP imaging with an inversion time of 300 ms and segmented inversion recovery turboFLASH sequences with optimized inversion recovery to null myocardial signal.

All images were analyzed by an image analyst with over 5 years of experience and all analyses were verified by a pediatric cardiologist with more than 10 years of experience. Ventricular function and volumes were calculated using Medis QMass (MedisSuite 2.1, Medis, Leiden, The Netherlands). The presence or absence of LGE was qualitatively assessed and localized using the standard 17-segment model^16^.

### Strain Analysis

We performed 4D CMR strain analysis using a custom-built graphical user interface in MATLAB (R2022b, Mathworks, Natick, Massachussetts, USA) as described previously^13^. In short, we first compiled short-axis cine CMR DICOM images into a volumetric data viewer that could reconstruct the axial, sagittal, and coronal planes of the heart. For each 4D image, the user then identified a centerline along the longitudinal axis of the left ventricle and tracked apical and basal movement along this axis.

Following this step, four equally spaced parallel short axis slices along this axis were identified. For each slice, the user tracked the movement of the endocardial and epicardial borders throughout the cardiac cycle in both the short axis and in one of three planes perpendicular to the short axis that corresponded approximately to conventional long-axis, two-chamber, and four-chamber views. This method amounts to tracking a structured set of 48 points throughout the myocardium in addition to basal and apical movement across a representative cardiac cycle. Following this step, we used spatial and temporal spline interpolation to create a dynamic 3D deformable mesh representing the left ventricle^13, 17, 18^. We then used this mesh to derive the following components of strain throughout the entire 3D mesh: circumferential strain (E_cc_), longitudinal strain (E_ll_), radial strain (E_rr_), and surface area strain (E_a_) in a Lagrangian reference frame as described previously^13^. From each strain component, we also derived regional average strain rates by measuring the slope of the strain curve in systole, early diastole, and late diastole, as described previously^13, 19^.

### Statistical Analysis

We assessed the distribution for each metric using a Shapiro-Wilk test (*p* < 0.05). Non-parametric statistics were used for data sets not following a normal distribution. When comparing two groups, we used t-test or Mann-Whitney test for parametric and non-parametric data respectively, and used the Holm-Sidak method for adjusting the p-value for multiple comparisons. When comparing multiple groups, we used an ANOVA with the Tukey-Kramer method for multiple comparisons for parametric data or the Kruskal-Wallis test and Dunn’s multiple comparison test for non-parametric data. For comparing categorical variables (i.e. LGE presence) we used a Fisher Exact Test.

## Results

Over the study period, we included 115 CMR studies in our analysis from n=40 patients. We imaged all of these patients using gadolinium contrast. LVEF, LGE status, demographic and clinical parameters in addition to other imaging variables were collected as outlined below in Table 1.

**Table 1:**
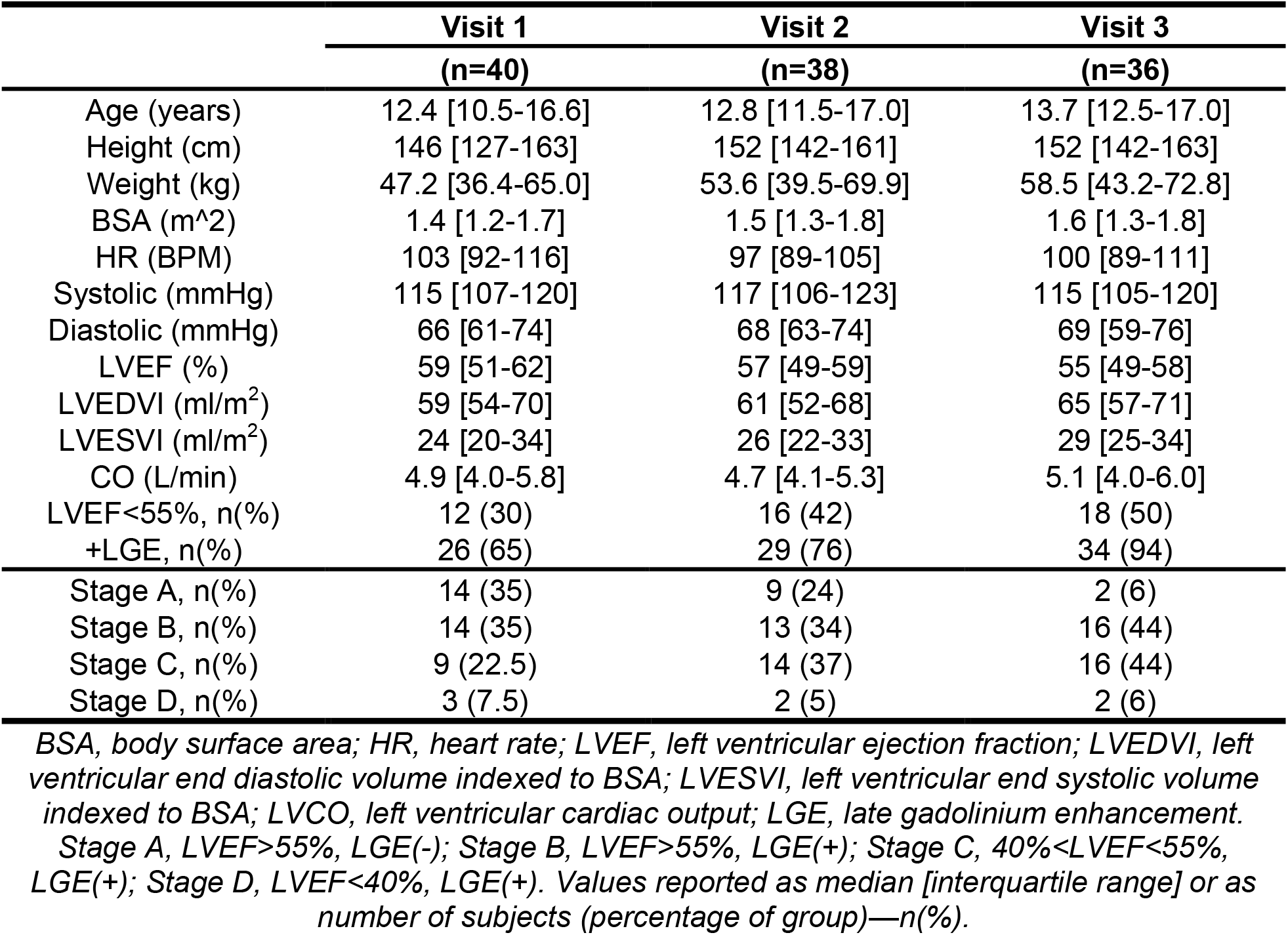
Clinical information obtained over three annual visits from patients with Duchenne muscular dystrophy-associated cardiomyopathy (DMD CMP).

### Late Gadolinium Enhancement

We measured the presence of LGE in each patient and found that the percentage of patients with LGE increased over time from 65% (n=26) at the initial visit to 76% (n=29) at +1 year and 94% (n=34) at +2 years. Additionally, the highest number of patients had findings of LGE in the basal and lateral segments of the heart with 81% (n=29) of patients having LGE in the basal inferolateral segment of the heart and 78% (n=28) in the basal anterolateral segment of the heart (Figure 2B).

**Figure 1:**
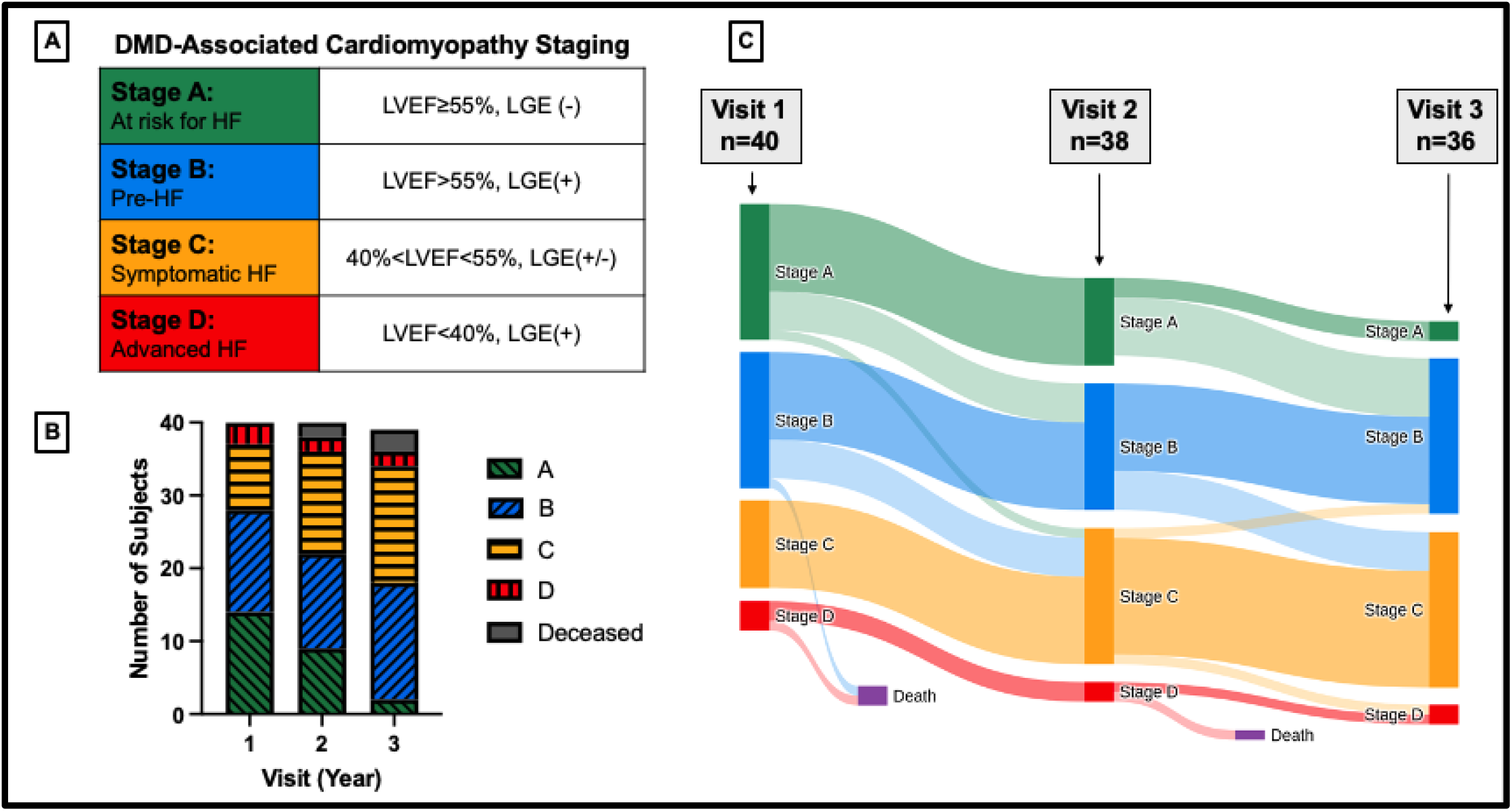
DMD CMP heart failure staging and cohort overview. A) Heart failure staging criteria. B) Stacked column chart depicting patient groups at visit 1 (initial visit), visit 2 (+1 year), and visit 3 (+2 years). C) Sankey diagram depicting heart failure stages across visits.

**Figure 2:**
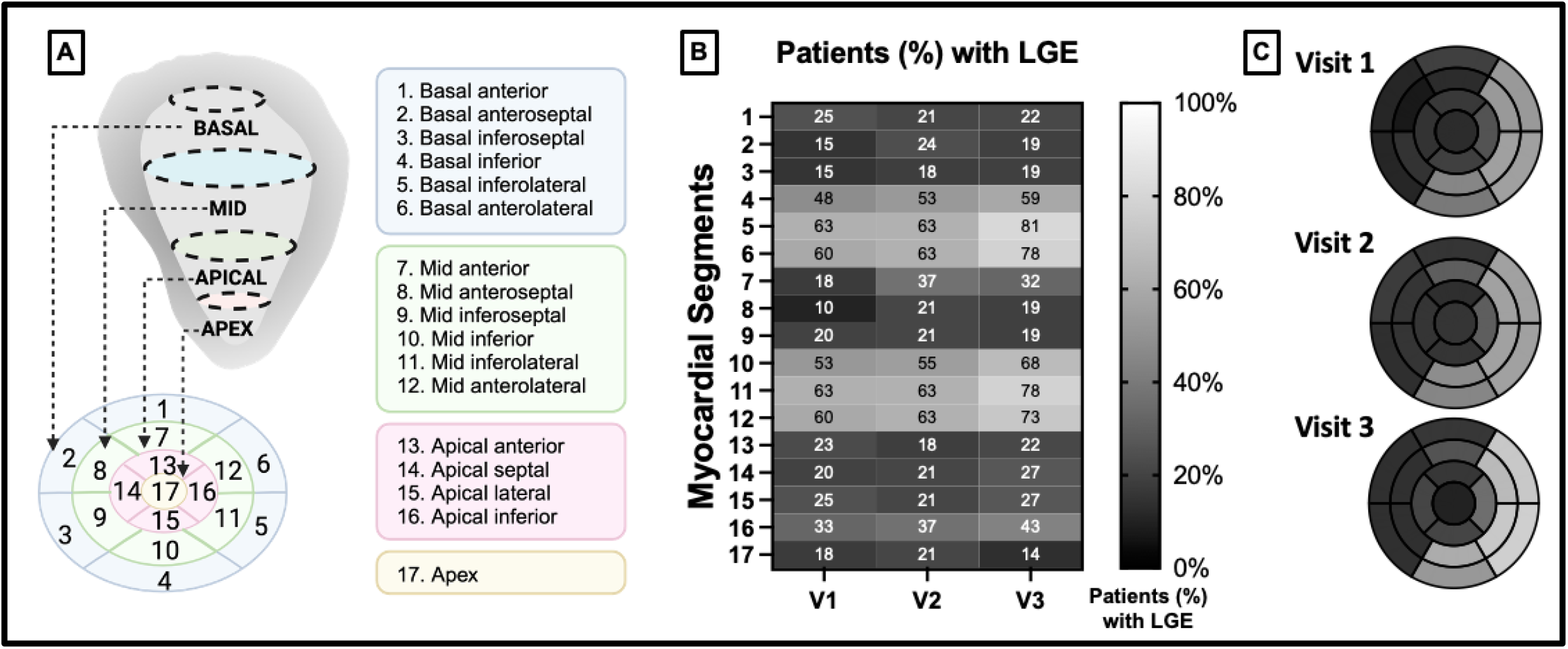
LGE presence increases over time in DMD, particularly in the basal and lateral regions of the heart. A) 17-segment polar plot map. B) Intensity plot depicting the percentage of patients with LGE findings for visit 1 (V1; initial visit; n=40), visit 2 (V2; +1 year; n=38), and visit 3 (V3; +2 years; n=36), with lighter shading depicting a higher percentage of LGE. C) 17-segment plot depicting the percentage of the patient cohort with LGE based on region, with lighter shading depicting a higher percentage of LGE.

### Strain Mapping and Progression

We calculated circumferential (E_cc_), longitudinal (E_ll_), radial (E_rr_), and surface area strain (E_a_) from 3D+time CMR images for each patient at the initial, second, and third visit. We plotted E_cc_ and E_ll_ using a Cartesian color map where the region of the heart is displayed on the y-axis and relative time in the cardiac cycle is depicted on the x-axis (Figure 3 A-D). We also plotted E_rr_ and E_a_ at peak systole using a polar plot (Figure 3 A-D). For many patients, we found striking qualitative differences in strain maps between visits indicating an overall decrease in strain magnitude over time (Figure 3).

**Figure 3:**
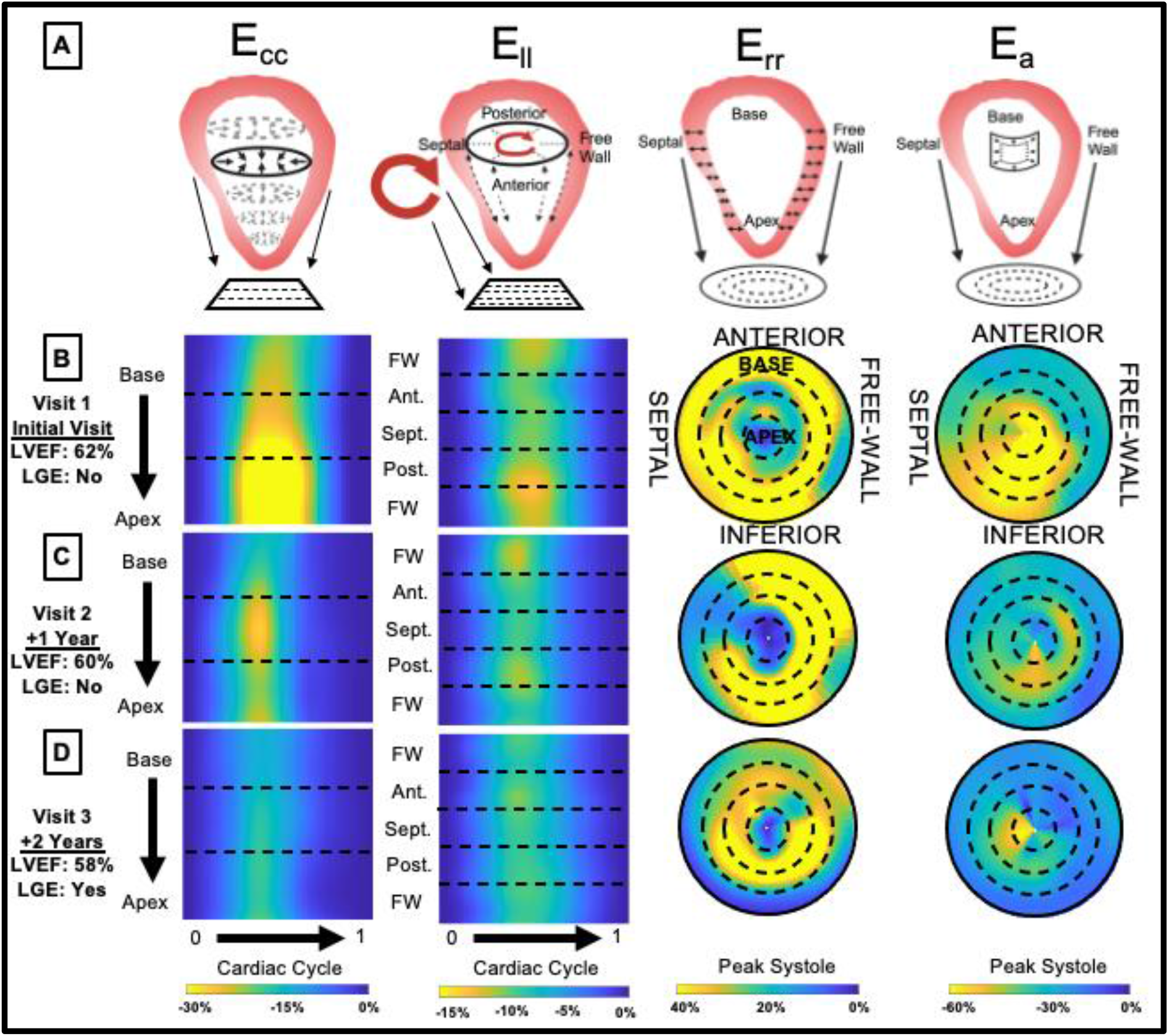
Regional 4D CMR strain metrics change over time in an individual patient. A) Graphical depiction of strain component calculation and graphical representation for circumferential (E_cc_), longitudinal (E_ll_), radial (E_rr_), and surface area strain (E_a_). B,C,D) Strain maps for an individual patient at the initial visit (B), 2^nd^ visit: +1 year (C), and 3^rd^ visit: +2 years (D).

Using the same strain mapping conventions, we also plotted composite group-averaged strain maps to evaluate averaged strain differences and patterns between groups with varying CMP severity (Figure 4). While there were no statistical difference between LVEF for Stage A (61.6±3.1%; mean±SD; n=14) and Stage B (61.5±5.2%; n=14; *p*=0.93), there were significant differences in global peak strain values between Stage A and Stage B. These differences included E_ll_ (−10.9±1.5, −8.6±1.4, *p*=0.002), E_rr_ (27±9.5, 18.2±6.3, *p*=0.038), and E_a_ (−31.9±3.4, −28±3.9, *p*=0.037). There were no significant differences in global peak strain between Stage B and Stage C, though differences approached significance for basal E_a_ peak strain (−25.2±2.8, −22.4±1.8, *p*=0.057). There were, however, significant differences in global peak strain between Stage C and Stage D for E_cc_ (−9.6±1.7, −5.9±2.4, *p*=0.017) and E_a_ (−24.9±1.9, −16.2±5.5, *p*=0.004). Additional regional peak differences are shown in Supplemental Table 1.

**Figure 4:**
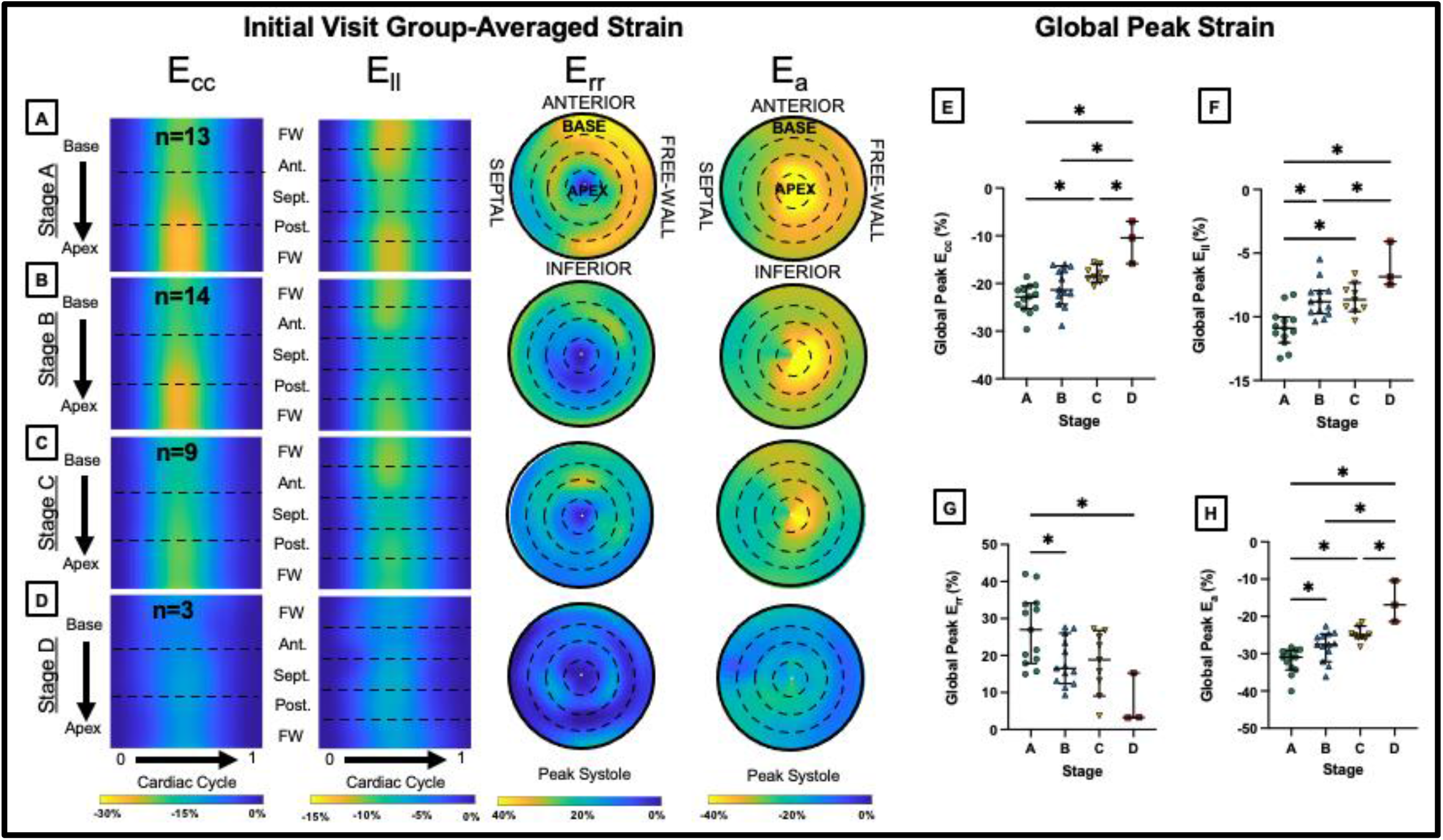
Regional 4D CMR strain metrics can differentiate between heart failure stages. A,B,C,D) Initial visit, group-averaged circumferential (E_cc_), longitudinal (E_ll_), radial (E_rr_), and surface area strain (E_a_) maps for Stage A (LVEF≥55%, no LGE), B (LVEF≥55%, with LGE), C (40%<LVEF<55%, with LGE), or D (LVEF<40%, with LGE). E,F,G,H) Global peak systolic strain values comparing heart failure stages at the initial visit for E_cc_ (E), E_ll_ (F), E_rr_ (G), and E_a_ (H). \**p*<0.05 adjusted for multiple comparisons.

### Strain Mapping and Progression

We further examined strain differences based on the progression of CMP from the initial visit as measured by the change in LVEF. As described in the methods, we grouped our patients into normal (LVEF≥55%) and abnormal (LVEF<55%) groups and further into slow (+2 years: ΔLVEF%≤5) or fast (+2 years: ΔLVEF%>5) progressors based on individual decrease in LVEF (Figure 5A). We were particularly interested in distinguishing progression differences between individuals with baseline normal function and those with baseline abnormal function. We found that at the initial visit, traditional functional metrics did not show statistical differences in the normal cohort between slow and fast progression including LVEF (*p*>0.99), age (*p*>0.99), and LGE status (*p*=0.704; Figure 5B,C). We saw similar trends in the abnormal cohort between slow and fast progression groups for LVEF (*p*=0.70) and age (*p*>0.99; Figure 5B,C). We did, however, note distinct qualitative differences in initial visit group-averaged strain maps between each progression group (Figure 5D-G) as well as in strain maps from representative individuals from each group (Supplemental Figures 1-4).

**Figure 5:**
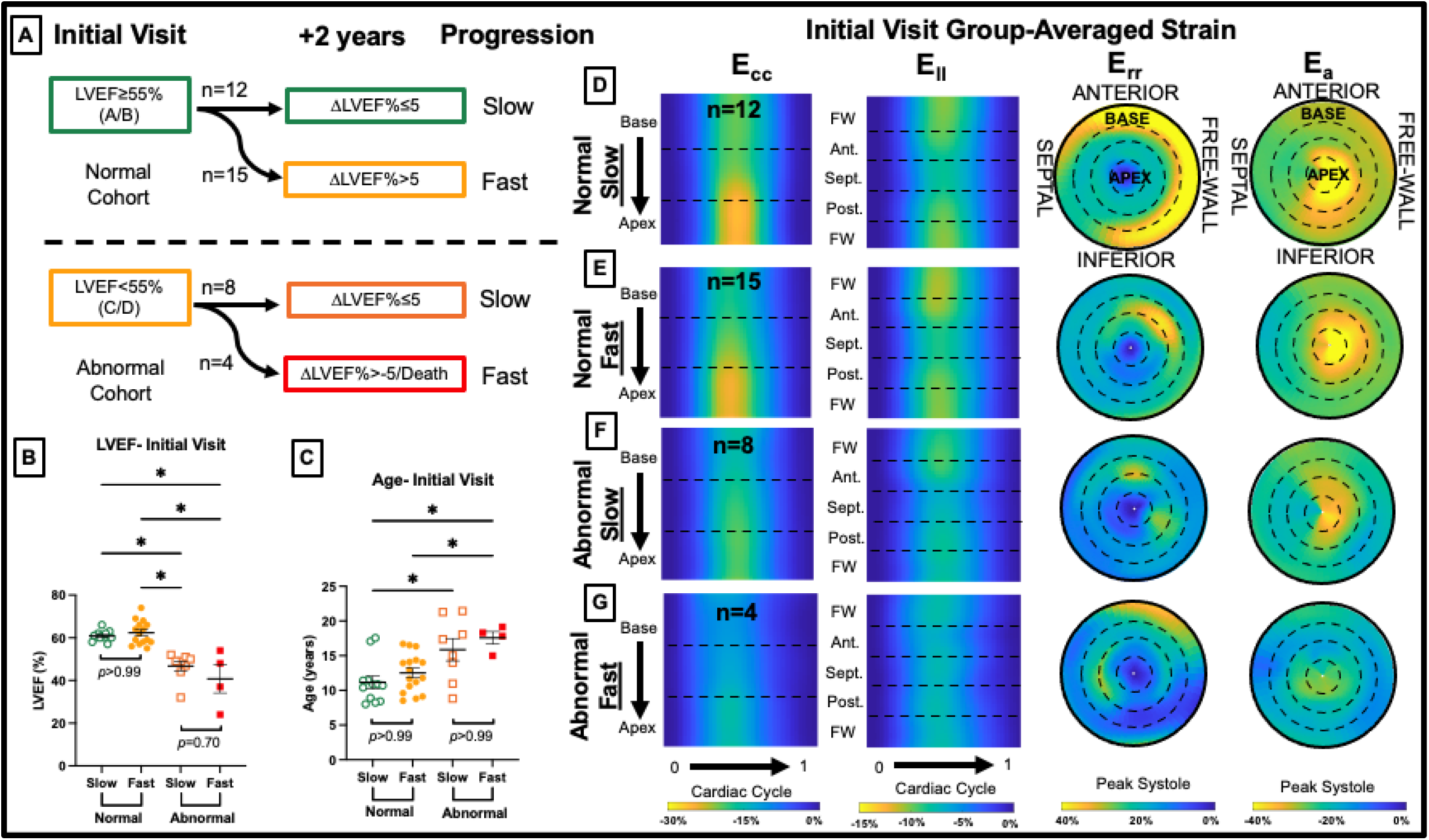
Standard metrics cannot differentiate between fast and slow progression; however, group-averaged strain maps reveal distinct qualitative differences between groups. A) Progression group descriptions for normal, slow progression (initial LVEF≥55% +2 years ΔLVEF%≤5), normal, fast progression (initial LVEF≥55% +2 years ΔLVEF%>5), abnormal, slow progression (initial LVEF<55% +2 years ΔLVEF%≤5), and abnormal, fast progression (initial LVEF<55% +2 years ΔLVEF%>5). B,C) LVEF comparison (B) and age comparison (C) at the initial visit between groups. D,E,F,G) Group averaged strain plots at the initial visit. \**p*<0.05 adjusted for multiple comparisons.

We also found some distinct quantitative differences in regional strain metrics between groups (Supplemental Tables 2 and 3). Patients with initially normal LVEF and fast progression had decreased peak systolic strain magnitude (Figure 6A) in the basal region only for E_cc_ (*p*=0.002), E_rr_ (*p*=0.001), and E_a_ (*p*=0.003; Figure 6B,C; Supplemental Table 2) compared to those with slow progression. There were no differences in the normal cohort between fast and slow progression for systolic strain rate. However, the fast progressors had decreased early diastolic strain rate magnitude for basal E_rr_ (*p*=0.001) and global E_rr_ (*p*=0.044). There were also differences in late diastolic strain rate for E_cc_ in the basal (*p*<0.001), mid-LV (*p*=0.026), and global (*p*=0.026) regions (Figure 6F) and for E_a_ in the basal (*p*=0.001), mid-LV (*p*=0.042), and global (*p*=0.042) regions (Figure 6G). Between fast and slow progressors in the abnormal cohort, there were no significant differences in peak systolic strain or systolic strain rate, however, there was a difference in early diastolic strain rate for E_ll_ in the anterior segment (*p*=0.048; Supplemental Table 3). Additionally, late diastolic strain rate magnitude was decreased in fast progressors for E_cc_ in the basal (*p=*0.045) and mid-LV (*p*=0.031) regions (Figure 6H) and for E_a_ in the basal (*p*=0.018), mid-LV (*p*=0.018) regions, and globally (*p*=0.018; Figure 6I).

**Figure 6:**
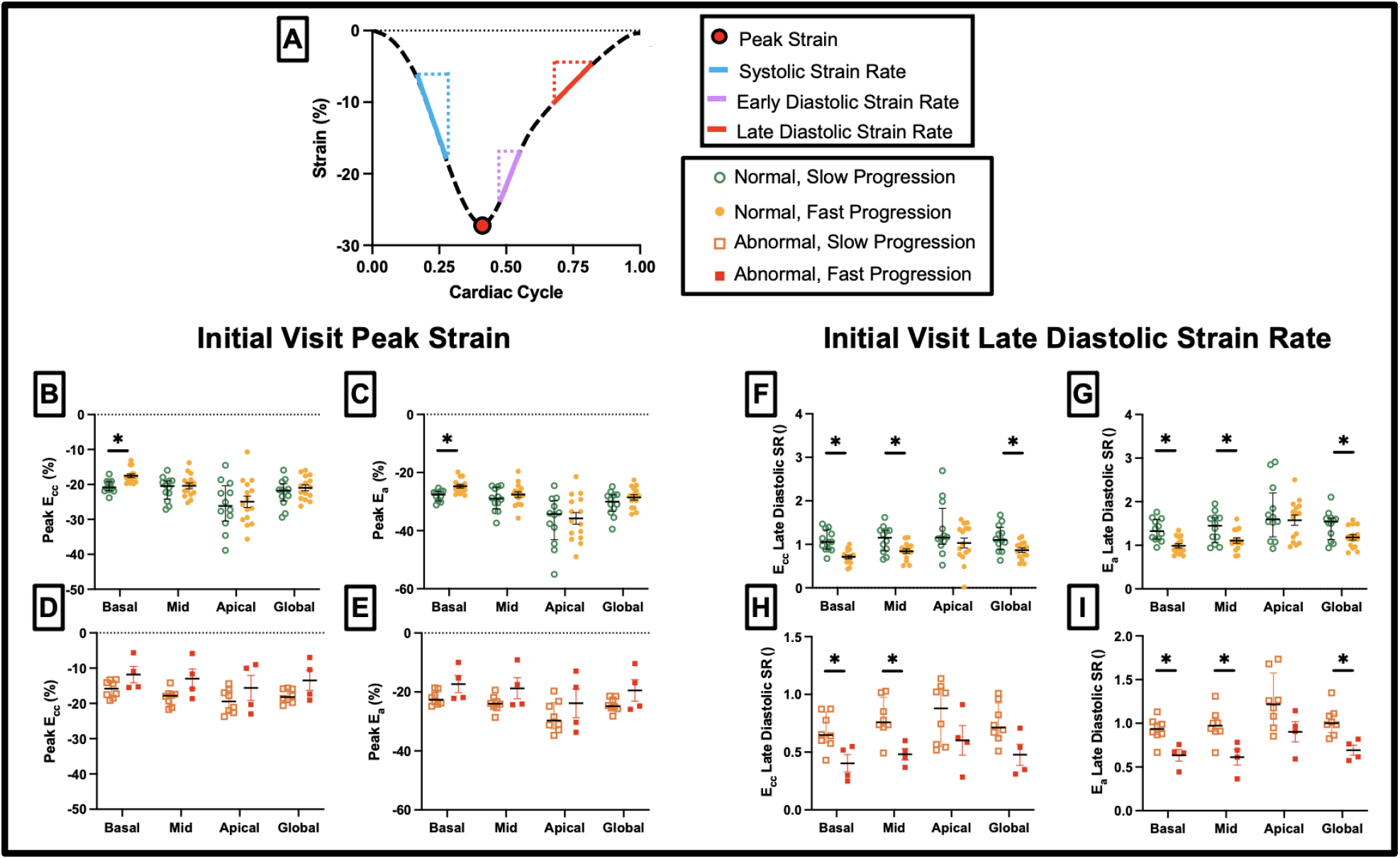
4D CMR regional peak strain and late diastolic strain rate can differentiate between slow and fast progression in both subclinical and clinical CMP. A) Strain curve depiction. B,C,D,E) Initial visit peak systolic strain showing differences between normal, slow progressors (initial LVEF≥55% +2 years ΔLVEF%≤5) and normal, fast progressors (initial LVEF≥55% +2 years ΔLVEF%>5) for E_cc_ (B) and E_a_ (C). Peak strain differences between abnormal, slow progressors (initial LVEF<55% +2 years ΔLVEF%≤5) and abnormal, fast progressors (initial LVEF<55% +2 years ΔLVEF%>5) are shown for E_cc_ (D) and E_a_ (E). F,G,H,I) Late diastolic strain rate differences are shown between fast and slow progression in the normal cohort for E_cc_ (F) and E_a_ (G) and between fast and slow progression in the abnormal cohort for E_cc_ (H) and E_a_ (I). \**p*<0.05 adjusted for multiple comparisons.

## Discussion

We report here that regional 4D CMR strain parameters are useful for stratifying patients based on both disease status and risk of progression. In this study, we used diagnostic CMR along with regional LGE to classify and phenotype a cohort of DMD patients followed annually over three visits. Further, we utilized a novel 4D CMR reconstruction and feature-tracking approach to derive and assess regional strain parameters. Finally, we used these strain-based parameters to evaluate differences in the rate of progression between patient groups prior to changes in LVEF or LGE. These findings and the metrics described here can be used to better describe the progression of CMP in a cohort of DMD patients followed longitudinally over time. This information is vital for improving the understanding and characteristics of this disease and justifies an increased need for larger, multi-center natural history studies in this patient population. It remains to be seen if 4D CMR strain parameters are modifiable or respond to treatment to delay CMP progression.

### Late Gadolinium Enhancement (LGE)

LGE is an important and early finding In DMD indicating the progression of CMP and fibrofatty replacement of cardiac muscle. In our cohort which had a wide range of CMP severity, we found that LGE presence increased over time from the initial and that the presence of LGE preceded patients having an LVEF<55%, a finding similar to other studies^20^. We also found that the greatest proportion of patients developed LGE in the basal and lateral portions of the left ventricle (Figure 2). At the initial visit, 25 out of the 26 patients with LGE had findings of LGE in the basal inferolateral region. There was also the greatest increase in the proportion of patients with LGE in this region. For example, there was an increase in the proportion of patients with LGE from 63% (n=25) to 81% (n=29) in the basal anterolateral region throughout the study. These findings are consistent with other studies that have shown regional differences in LGE presence in DMD patients^15, 21^ and offer further evidence for the importance of regional assessment.

This regional variation in LGE accumulation might be partially explained by the differences in stress within the left ventricle governed by the Law of Laplace. This law describes how such factors as pressure, local curvature, and increased myocardial thickness contribute to stress which is relatively higher in the basal segments of the heart^22, 23^. Thus, LGE accumulation in the base of the heart suggests strain parameters in the basal region may be useful for differentiating disease severity and progression.

### Heart Failure Staging

In previous work, we showed how 4D CMR regional strain parameters could strongly differentiate between DMD and healthy control patients^13^. In this study, we demonstrated that global peak strain could also differentiate between patients in Stage A (LVEF≥55%, no LGE) and Stage B (LVEF≥55% with LGE; Figure 4). This is an important finding because traditional metrics, such as LVEF, do not differentiate between these groups. This suggests that 3D+time strain metrics may be a useful non-contrast method for identifying the presence of LGE and/or fibrosis. Non-contrast methods to characterize CMP onset and progression could decrease scan time, reduce the need for contrast (LGE), and expand the number of DMD patients who could undergo CMR^15^. In summary, these data suggest that 4D CMR strain might be used for stratifying patient risk, increasing access to imaging, and guiding clinical management.

### Strain Mapping and Progression

One of the most clinically challenging aspects of DMD CMP care is understanding the risk profile of patients. By examining DMD patients with a range of CMP severity, we were able to identify patients who were more likely to experience rapid progression over the course of two years. We also distinguished between patient groups who initially had subclinical CMP (LVEF≥55%) versus those who had evidence of CMP (LVEF<55%). We made this initial distinction as it represents two clinically distinct groups with differences in their short-term natural history that might be of interest to clinical providers. Interestingly, we found that conventional metrics of LVEF, age, or LGE status could not differentiate between fast or slow progression in overt or subclinical CMP. We did find, however, that regional 4D CMR strain parameters could make this distinction (Figure 6, Supplemental Table 2,3), suggesting this approach may be better for detecting early changes in cardiac function.

The strongest differentiators for CMP progression in patients with normal function were peak systolic basal E_cc_, E_a_, and E_rr_ strain, early diastolic basal E_rr_ strain rate, late diastolic basal E_cc_ rate, and late diastolic basal E_a_ strain rate. The strongest differentiators for CMP progression in patients with abnormal function were regional late diastolic E_cc_ and E_a_ strain rates. These results indicate that, just as we observed in the LGE pattern, changes in the kinematics of the basal region of the heart may be an early indication of disease severity and potential for progression. These results are consistent with the findings of other studies using 3D strain in DMD. Siddiqui *et al.* showed previously that for DMD patients with an initial LVEF≥55% reductions in global 3D E_cc_ and E_rr_ could be indicative of a >10% decline in LVEF up to 32 months later^24^.

Another important finding of this study is the additive value of using regional diastolic strain rate as a metric for differentiating patient progression. Others have shown that abnormalities in diastolic function may be an early indication of CMP onset^25^. To our knowledge, this is the first study that describes the use of regional diastolic strain rate from 4D CMR as an early metric for CMP progression in DMD. Impaired diastolic relaxation in these patients may be an early sign of ventricular stiffening from fibrofatty replacement of cardiomyocytes and impaired kinematics. Despite this progress, further focus and study on both systolic and diastolic strain-based metrics in this population is merited.

### Strengths and Limitations

This study has several strengths and limitations. The analysis currently requires a post-processing approach using a custom image analysis code for 4D CMR analysis. Incorporation of this method or similar approach in combination with emerging artificial intelligence (AI) assisted feature tracking would allow this kind of analysis to be automated and performed within a reasonable clinical image analysis workflow. Additionally, several of the short-axis stacks were shifted relative to others in the 3D stack due to breathing and motion artifacts. This could have been partially due to the young age of the patients, who may be unable to lie still during imaging for long periods of time. Using contextual information from other slices, we can still evaluate these images, but there was one dataset that we were unable to reconstruct in three dimensions due to these artifacts. AI-assisted motion correction and CMR sequences that do not rely on breath-holding could overcome these artifacts in the future. Further, this is a cohort followed over three annual imaging sessions with a relatively wide range of CMP severity. Though our sample size is relatively high for a rare disease, the smaller sample size limits the power of our study, in particular when performing subgroup analysis. Because of the limited sample size, we also did not adjust for medication effects on our cohort, though all patients were treated at the same clinic with standard of care and a proactive treatment approach. Therefore, the range and differences in progression in our cohort are likely very similar to what a clinician would be seeing in practice. Future studies with larger group sizes should adjudicate treatment affects.

Strengths of this study include the longitudinal follow-up of patients, comprehensive regional 4D CMR strain analysis, and the study design allowing us to evaluate the rate of progression among our patients. The longitudinal analysis of patients allowed us to evaluate both individual patient and group averaged progression as well as changes over time. Additionally, our novel 4D CMR feature tracking software allowed us to assess not only global function but also detailed spatial and temporal information for each strain component. Here we also describe the use and value of surface area strain (E_a_) which is a parameter unique to gated 3D imaging as it is a measure of 3D surface deformation. To our knowledge, this is the first description of 4D CMR strain parameters for differentiation of disease severity and both subclinical and clinical rates of progression.

### Conclusion

In this study, we described the value of 4D CMR imaging metrics for differentiating both CMP disease severity and rate of progression. We showed that basal region peak strain and late diastolic strain rate for E_cc_ and E_a_ were correlated with the rate of cardiac disease progression in DMD patients. Further studies assessing the natural history and progression of DMD CMP using these techniques could help assess individual patient risk profiles and guide treatment to improve clinical outcomes.

## Data Availability

Data referred to in the manuscript is available from the corresponding author upon reasonable request.

## Acknowledgments

The authors acknowledge senior imaging research specialist Kristen George-Durrett for her help and expertise in image data management.

## Supplemental Tables and Figures

**Supplemental Figure 1:**
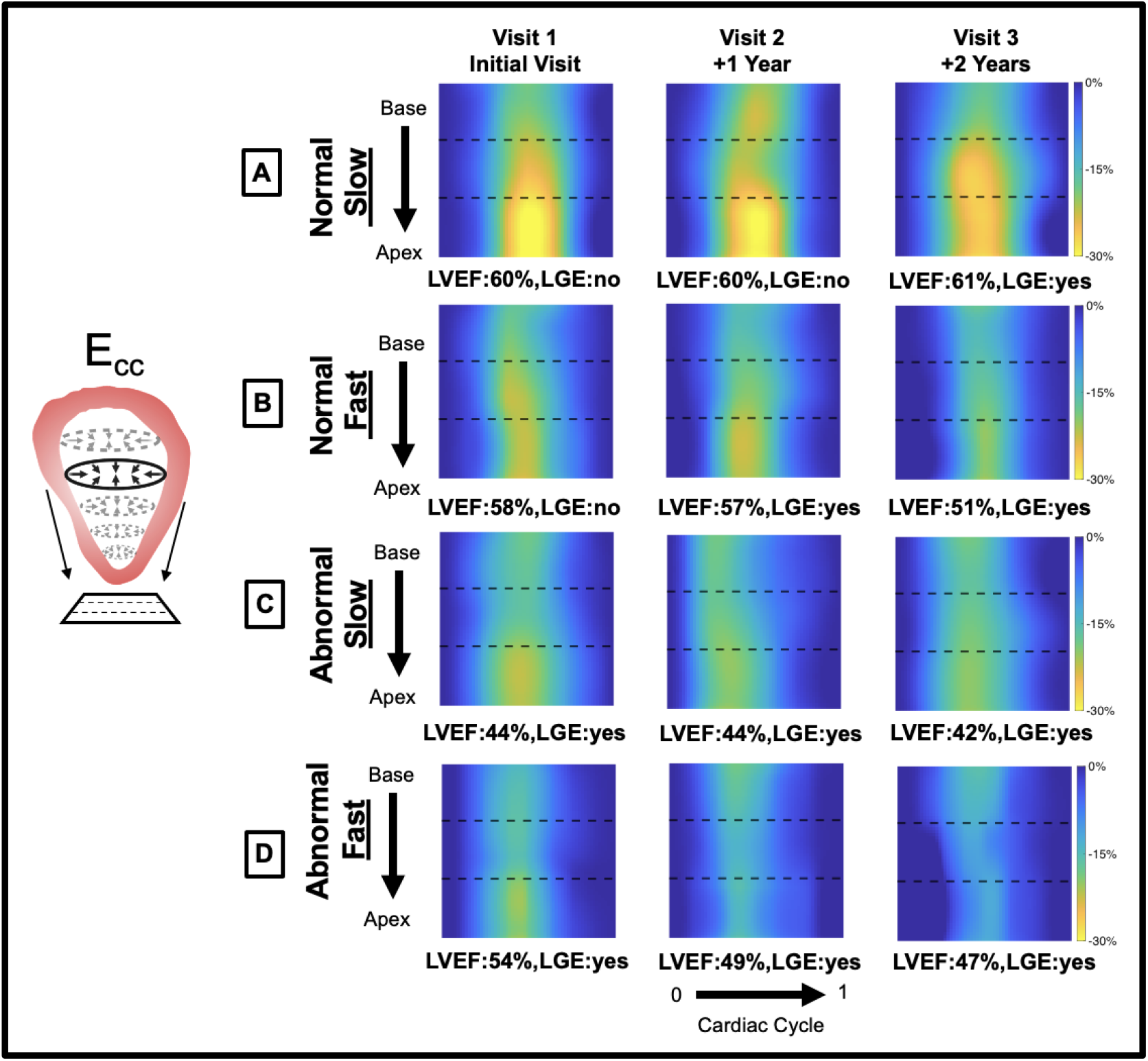
Circumferential strain (E_cc_) progression. A,B,C,D) E_cc_ Cartesian maps showing differences between a normal, slow progressor (A; initial LVEF≥55% +2 years ΔLVEF%≤5), normal, fast progressor (B; initial LVEF≥55% +2 years ΔLVEF%>5), abnormal, slow progressor (C; initial LVEF<55% +2 years ΔLVEF%≤5), abnormal, fast progressor (D; initial LVEF<55%, +2 years ΔLVEF%>5) over the course of three annual visits.

**Supplemental Figure 2:**
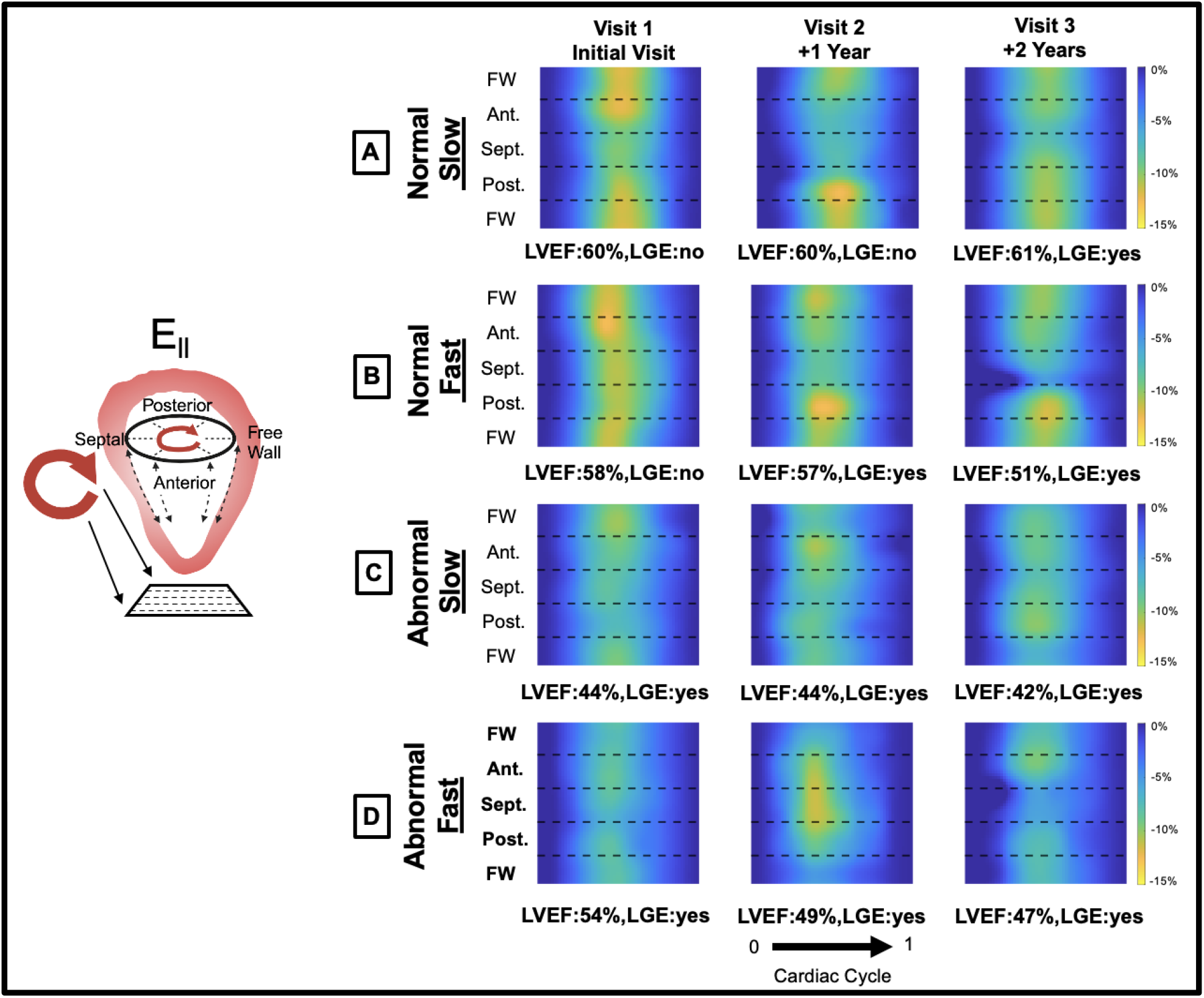
Longitudinal strain (E_ll_) progression. A,B,C,D) E_ll_ Cartesian maps showing differences between a normal, slow progressor (A; initial LVEF≥55% +2 years ΔLVEF%≤5), normal, fast progressor (B; initial LVEF≥55% +2 years ΔLVEF%>5), abnormal, slow progressor (C; initial LVEF<55% +2 years ΔLVEF%≤5), abnormal, fast progressor (D; initial LVEF<55%, +2 years ΔLVEF%>5) over the course of three annual visits.

**Supplemental Figure 3:**
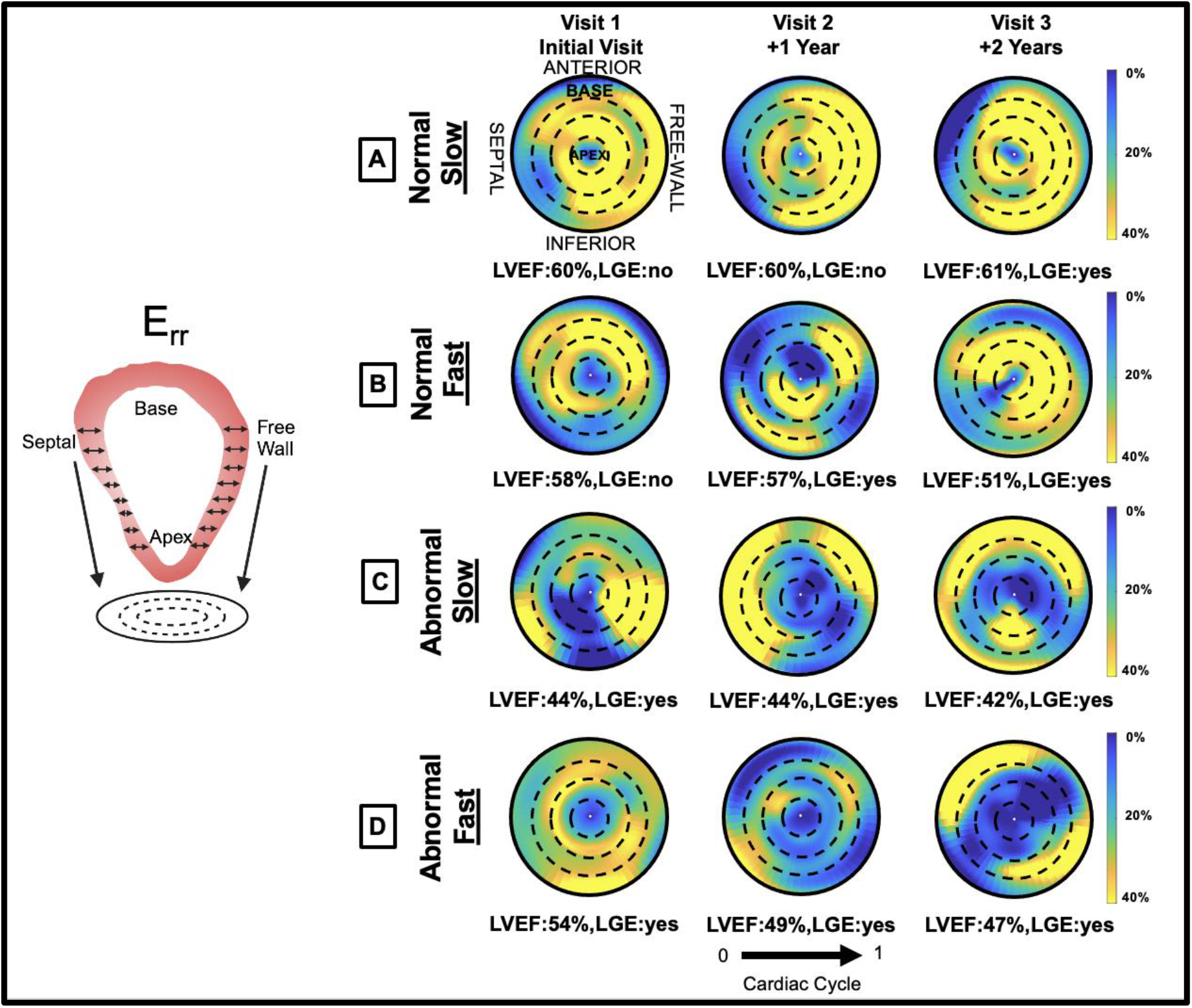
Radial strain (E_rr_) progression. A,B,C,D) E_rr_ polar strain maps at peak systole showing differences between a normal, slow progressor (A; initial LVEF≥55% +2 years ΔLVEF%≤5), normal, fast progressor (B; initial LVEF≥55% +2 years ΔLVEF%>5), abnormal, slow progressor (C; initial LVEF<55% +2 years ΔLVEF%≤5), abnormal, fast progressor (D; initial LVEF<55%, +2 years ΔLVEF%>5) over the course of three annual visits.

**Supplemental Figure 4:**
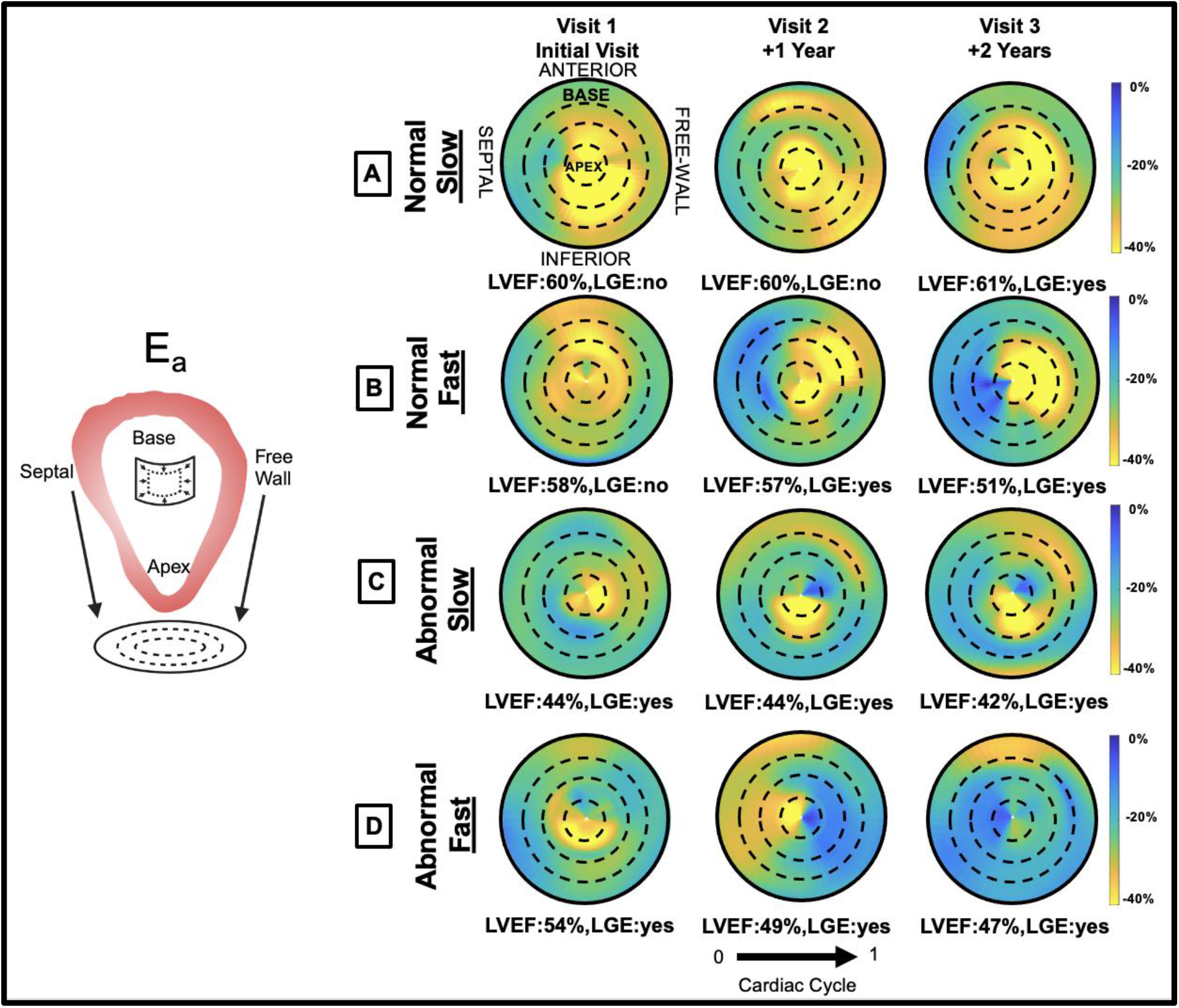
Surface area strain (E_a_) progression. A,B,C,D) E_a_ polar strain maps at peak systole showing differences between a normal, slow progressor (A; initial LVEF≥55% +2 years ΔLVEF%≤5), normal, fast progressor (B; initial LVEF≥55% +2 years ΔLVEF%>5), abnormal, slow progressor (C; initial LVEF<55% +2 years ΔLVEF%≤5), abnormal, fast progressor (D; initial LVEF<55%, +2 years ΔLVEF%>5) over the course of three annual visits.

**Supplemental Table 1:**
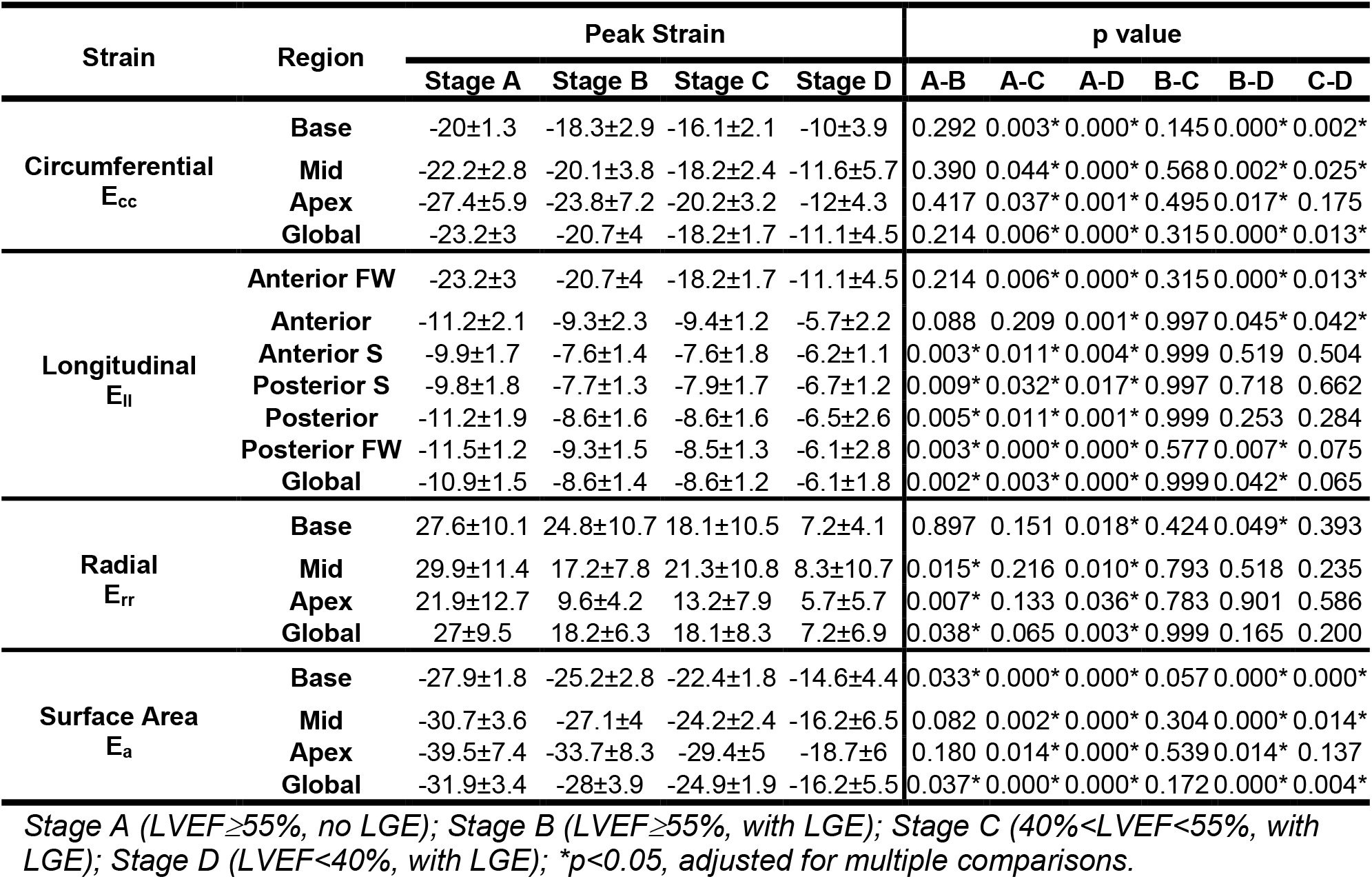
Regional 4D CMR peak systolic strain differences between heart failure stages.

| Strain | Region | Peak Strain |  |  |  | p value |  |  |  |  |  |
| --- | --- | --- | --- | --- | --- | --- | --- | --- | --- | --- | --- |
|  |  | Stage A | Stage B | Stage C | Stage D | A-B | A-C | A-D | B-C | B-D | C-D |
| Circumferential<br>$E_{cc}$ | Base | -20±1.3 | -18.3±2.9 | -16.1±2.1 | -10±3.9 | 0.292 | 0.003* | 0.000* | 0.145 | 0.000* | 0.002* |
|  | Mid | -22.2±2.8 | -20.1±3.8 | -18.2±2.4 | -11.6±5.7 | 0.390 | 0.044* | 0.000* | 0.568 | 0.002* | 0.025* |
|  | Apex | -27.4±5.9 | -23.8±7.2 | -20.2±3.2 | -12±4.3 | 0.417 | 0.037* | 0.001* | 0.495 | 0.017* | 0.175 |
|  | Global | -23.2±3 | -20.7±4 | -18.2±1.7 | -11.1±4.5 | 0.214 | 0.006* | 0.000* | 0.315 | 0.000* | 0.013* |
| Longitudinal<br>$E_{ll}$ | Anterior FW | -23.2±3 | -20.7±4 | -18.2±1.7 | -11.1±4.5 | 0.214 | 0.006* | 0.000* | 0.315 | 0.000* | 0.013* |
|  | Anterior | -11.2±2.1 | -9.3±2.3 | -9.4±1.2 | -5.7±2.2 | 0.088 | 0.209 | 0.001* | 0.997 | 0.045* | 0.042* |
|  | Anterior S | -9.9±1.7 | -7.6±1.4 | -7.6±1.8 | -6.2±1.1 | 0.003* | 0.011* | 0.004* | 0.999 | 0.519 | 0.504 |
|  | Posterior S | -9.8±1.8 | -7.7±1.3 | -7.9±1.7 | -6.7±1.2 | 0.009* | 0.032* | 0.017* | 0.997 | 0.718 | 0.662 |
|  | Posterior | -11.2±1.9 | -8.6±1.6 | -8.6±1.6 | -6.5±2.6 | 0.005* | 0.011* | 0.001* | 0.999 | 0.253 | 0.284 |
|  | Posterior FW | -11.5±1.2 | -9.3±1.5 | -8.5±1.3 | -6.1±2.8 | 0.003* | 0.000* | 0.000* | 0.577 | 0.007* | 0.075 |
|  | Global | -10.9±1.5 | -8.6±1.4 | -8.6±1.2 | -6.1±1.8 | 0.002* | 0.003* | 0.000* | 0.999 | 0.042* | 0.065 |
| Radial<br>$E_{rr}$ | Base | 27.6±10.1 | 24.8±10.7 | 18.1±10.5 | 7.2±4.1 | 0.897 | 0.151 | 0.018* | 0.424 | 0.049* | 0.393 |
|  | Mid | 29.9±11.4 | 17.2±7.8 | 21.3±10.8 | 8.3±10.7 | 0.015* | 0.216 | 0.010* | 0.793 | 0.518 | 0.235 |
|  | Apex | 21.9±12.7 | 9.6±4.2 | 13.2±7.9 | 5.7±5.7 | 0.007* | 0.133 | 0.036* | 0.783 | 0.901 | 0.586 |
|  | Global | 27±9.5 | 18.2±6.3 | 18.1±8.3 | 7.2±6.9 | 0.038* | 0.065 | 0.003* | 0.999 | 0.165 | 0.200 |
| Surface Area<br>$E_a$ | Base | -27.9±1.8 | -25.2±2.8 | -22.4±1.8 | -14.6±4.4 | 0.033* | 0.000* | 0.000* | 0.057 | 0.000* | 0.000* |
|  | Mid | -30.7±3.6 | -27.1±4 | -24.2±2.4 | -16.2±6.5 | 0.082 | 0.002* | 0.000* | 0.304 | 0.000* | 0.014* |
|  | Apex | -39.5±7.4 | -33.7±8.3 | -29.4±5 | -18.7±6 | 0.180 | 0.014* | 0.000* | 0.539 | 0.014* | 0.137 |
|  | Global | -31.9±3.4 | -28±3.9 | -24.9±1.9 | -16.2±5.5 | 0.037* | 0.000* | 0.000* | 0.172 | 0.000* | 0.004* |

Stage A (LVEF≥55%, no LGE); Stage B (LVEF≥55%, with LGE); Stage C (40%<LVEF<55%, with LGE); Stage D (LVEF<40%, with LGE); \*p<0.05, adjusted for multiple comparisons.

**Supplemental Table 2:** Regional strain differences between cardiomyopathy groups with normal LVEF:

| Strain | Region | Peak |  |  | Systolic Strain Rate |  |  | Early Diastolic Strain Rate |  |  | Late Diastolic Strain Rate |  |  |
| --- | --- | --- | --- | --- | --- | --- | --- | --- | --- | --- | --- | --- | --- |
|  |  | Group 1 | Group 2 | p | Group 1 | Group 2 | p | Group 1 | Group 2 | p | Group 1 | Group 2 | p |
| Circumferential<br>$E_{cc}$ | Base | -20.5±1.8 | -17.5±2.1 | 0.002* | -1.19±0.3 | -1.04±0.1 | 0.348 | 1.05±0.3 | 0.85±0.2 | 0.193 | 1.08±0.2 | 0.71±0.2 | 0.000* |
|  | Mid | -21.3±3.4 | -20.4±3.2 | 0.745 | -1.36±0.5 | -1.24±0.2 | 0.365 | 1.16±0.4 | 0.99±0.2 | 0.23 | 1.11±0.3 | 0.84±0.2 | 0.026* |
|  | Apex | -25.9±6.9 | -25±6.4 | 0.745 | -1.78±0.6 | -1.54±0.4 | 0.364 | 1.57±0.6 | 1.32±0.4 | 0.23 | 1.35±0.6 | 1.03±0.4 | 0.134 |
|  | Global | -22.5±3.8 | -21±3.3 | 0.629 | -1.51±0.3 | -1.25±0.2 | 0.075 | 1.25±0.4 | 1.03±0.2 | 0.208 | 1.14±0.3 | 0.87±0.2 | 0.026* |
| Longitudinal<br>$E_{ll}$ | Anterior FW | -10.2±1.8 | -10.9±2 | 0.956 | -0.57±0.1 | -0.62±0.1 | 0.836 | 0.48±0.1 | 0.5±0.1 | 0.997 | 0.46±0.1 | 0.4±0.1 | 0.775 |
|  | Anterior | -9.7±2.7 | -10.7±2.2 | 0.929 | -0.56±0.2 | -0.61±0.1 | 0.945 | 0.45±0.1 | 0.45±0.1 | 0.999 | 0.4±0.1 | 0.42±0.1 | 0.976 |
|  | Anterior S | -8.4±2.1 | -8.9±1.8 | 0.979 | -0.47±0.1 | -0.5±0.1 | 0.945 | 0.36±0.1 | 0.37±0.1 | 0.999 | 0.37±0.1 | 0.35±0.1 | 0.976 |
|  | Posterior S | -8.5±1.8 | -8.8±1.9 | 0.979 | -0.48±0.1 | -0.51±0.1 | 0.945 | 0.37±0.1 | 0.36±0.1 | 0.999 | 0.37±0.1 | 0.34±0.1 | 0.976 |
|  | Posterior | -9.7±2.1 | -9.9±2.2 | 0.979 | -0.53±0.1 | -0.58±0.2 | 0.945 | 0.45±0.1 | 0.42±0.1 | 0.959 | 0.39±0.1 | 0.38±0.1 | 0.976 |
|  | Posterior FW | -10.3±1.8 | -10.3±1.6 | 0.979 | -0.58±0.1 | -0.58±0.1 | 0.945 | 0.5±0.1 | 0.48±0.1 | 0.999 | 0.46±0.1 | 0.38±0.1 | 0.309 |
|  | Global | -9.5±1.9 | -9.9±1.7 | 0.979 | -0.53±0.1 | -0.57±0.1 | 0.945 | 0.43±0.1 | 0.42±0.1 | 0.999 | 0.41±0.1 | 0.37±0.1 | 0.929 |
| Radial<br>$E_{rr}$ | Base | 34.6±10.2 | 20.2±6.9 | 0.001* | 1.77±0.9 | 1.03±0.5 | 0.054 | -2.65±0.7 | -1.5±0.7 | 0.001* | -1.29±1.1 | -0.63±0.5 | 0.204 |
|  | Mid | 23.2±7.5 | 22.3±15 | 0.979 | 1.02±0.4 | 1.38±1.2 | 0.695 | -1.96±0.6 | -1.48±1 | 0.256 | -0.58±0.8 | -0.93±0.9 | 0.672 |
|  | Apex | 15.6±9.2 | 15±12.7 | 0.979 | 0.75±0.8 | 0.86±0.8 | 0.88 | -1.31±0.8 | -1.12±1 | 0.596 | -0.5±0.8 | -0.58±1.1 | 0.935 |
|  | Global | 25.5±6.6 | 19.7±10.5 | 0.286 | 1.23±0.5 | 1.12±0.7 | 0.88 | -2.06±0.4 | -1.4±0.8 | 0.044* | -0.82±0.8 | -0.73±0.7 | 0.935 |
| Surface Area<br>$E_a$ | Base | -28±1.8 | -24.7±2.4 | 0.003* | -1.63±0.2 | -1.46±0.2 | 0.117 | 1.39±0.2 | 1.19±0.2 | 0.131 | 1.35±0.3 | 0.99±0.2 | 0.001* |
|  | Mid | -29.2±4.1 | -27.6±3.8 | 0.509 | -1.84±0.3 | -1.66±0.2 | 0.199 | 1.47±0.4 | 1.29±0.2 | 0.222 | 1.4±0.3 | 1.11±0.2 | 0.042* |
|  | Apex | -36.6±8.6 | -35.8±7.7 | 0.802 | -2.46±0.7 | -2.21±0.4 | 0.299 | 2.16±0.8 | 1.8±0.4 | 0.222 | 1.75±0.6 | 1.58±0.5 | 0.431 |
|  | Global | -30.6±4.1 | -28.6±3.5 | 0.452 | -1.92±0.3 | -1.72±0.2 | 0.199 | 1.61±0.4 | 1.38±0.2 | 0.152 | 1.47±0.3 | 1.18±0.2 | 0.042* |

Group 1: normal, slow progression (initial LVEF≥55%, +2 years ΔLVEF%≤5) and Group 2: normal, fast progression (initial LVEF≥55%, +2 years ΔLVEF%≤5); \* $p$ <0.05, adjusted for multiple comparisons.

**Supplemental Table 3:** Regional strain differences between DMD cardiomyopathy groups with abnormal LVEF.

| Strain | Region | Peak |  |  | Systolic Strain Rate |  |  | Early Diastolic Strain Rate |  |  | Late Diastolic Strain Rate |  |  |
| --- | --- | --- | --- | --- | --- | --- | --- | --- | --- | --- | --- | --- | --- |
|  |  | Group 3 | Group 4 | p | Group 3 | Group 4 | p | Group 3 | Group 4 | p | Group 3 | Group 4 | p |
| Circumferential<br>$E_{cc}$ | Base | -15.9±2.4 | -11.8±4.6 | 0.185 | -0.88±0.2 | -0.78±0.4 | 0.547 | 0.88±0.2 | 0.57±0.2 | 0.096 | 0.68±0.2 | 0.4±0.2 | 0.045* |
|  | Mid | -18.3±2.4 | -13±5.6 | 0.145 | -1.06±0.2 | -0.73±0.3 | 0.148 | 0.97±0.3 | 0.7±0.3 | 0.258 | 0.79±0.2 | 0.48±0.1 | 0.031* |
|  | Apex | -19.4±3.4 | -15.6±7.1 | 0.233 | -1.16±0.3 | -0.88±0.4 | 0.385 | 1.15±0.4 | 0.73±0.4 | 0.258 | 0.83±0.3 | 0.6±0.3 | 0.188 |
|  | Global | -17.9±1.9 | -13.5±5.7 | 0.185 | -1.03±0.2 | -0.79±0.3 | 0.334 | 1±0.2 | 0.66±0.3 | 0.148 | 0.75±0.2 | 0.48±0.2 | 0.053 |
| Longitudinal<br>$E_{ll}$ | Anterior FW | -9.1±1.9 | -7.8±3.4 | 0.951 | -0.49±0.1 | -0.41±0.2 | 0.942 | 0.44±0.2 | 0.34±0.1 | 0.867 | 0.36±0.1 | 0.28±0.1 | 0.489 |
|  | Anterior | -9.2±1.3 | -7.1±3.1 | 0.57 | -0.48±0.1 | -0.4±0.2 | 0.919 | 0.45±0.1 | 0.29±0.1 | 0.048* | 0.35±0.1 | 0.27±0.1 | 0.489 |
|  | Anterior S | -7.2±1.6 | -7.5±2.1 | 0.97 | -0.39±0.1 | -0.42±0.1 | 0.991 | 0.34±0.1 | 0.34±0.1 | 0.997 | 0.28±0.1 | 0.28±0.1 | 0.957 |
|  | Posterior S | -7.6±1.5 | -7.4±2 | 0.97 | -0.42±0.1 | -0.43±0.1 | 0.991 | 0.35±0.1 | 0.38±0.1 | 0.984 | 0.31±0.1 | 0.27±0.1 | 0.629 |
|  | Posterior | -8.3±1.8 | -7.6±2.7 | 0.97 | -0.45±0.1 | -0.43±0.2 | 0.991 | 0.38±0.1 | 0.35±0.1 | 0.984 | 0.34±0.1 | 0.24±0.1 | 0.452 |
|  | Posterior FW | -8.1±1.5 | -7.6±2.9 | 0.97 | -0.42±0.1 | -0.4±0.2 | 0.991 | 0.36±0.1 | 0.33±0.1 | 0.984 | 0.34±0.1 | 0.26±0.1 | 0.452 |
|  | Global | -8.2±1.2 | -7.4±2.6 | 0.961 | -0.44±0.1 | -0.4±0.1 | 0.99 | 0.38±0.1 | 0.33±0.1 | 0.897 | 0.32±0 | 0.24±0.1 | 0.397 |
| Radial<br>$E_{rr}$ | Base | 14.9±9.7 | 16.2±13.1 | 0.999 | 0.59±0.7 | 2.06±1.4 | 0.129 | -1.18±1 | -1.78±1.4 | 0.873 | -0.5±0.7 | -0.49±0.4 | 0.967 |
|  | Mid | 18.6±9.7 | 16.8±17 | 0.999 | 0.78±0.7 | 1.39±1.2 | 0.491 | -1.51±0.7 | -1.91±1.2 | 0.873 | -0.41±0.7 | -0.7±1 | 0.958 |
|  | Apex | 11.5±7.4 | 10.9±10.1 | 0.999 | 0.47±0.5 | 0.62±0.6 | 0.66 | -0.94±0.6 | -0.95±0.7 | 0.971 | -0.19±0.5 | -0.4±0.6 | 0.958 |
|  | Global | 15.5±7 | 15.1±13.7 | 0.999 | 0.63±0.6 | 1.45±1 | 0.261 | -1.24±0.6 | -1.62±1.1 | 0.873 | -0.39±0.6 | -0.55±0.7 | 0.958 |
| Surface Area<br>$E_a$ | Base | -22.1±2.4 | -17.3±5.8 | 0.238 | -1.17±0.1 | -1.13±0.5 | 0.843 | 1.19±0.2 | 0.88±0.2 | 0.076 | 0.93±0.1 | 0.63±0.1 | 0.018* |
|  | Mid | -23.9±2.7 | -18.8±7.2 | 0.25 | -1.34±0.1 | -1.09±0.5 | 0.605 | 1.29±0.2 | 0.98±0.2 | 0.088 | 0.98±0.2 | 0.61±0.2 | 0.018* |
|  | Apex | -28.2±5.2 | -23.8±9.8 | 0.323 | -1.62±0.3 | -1.42±0.5 | 0.753 | 1.64±0.5 | 1.28±0.5 | 0.272 | 1.25±0.3 | 0.9±0.2 | 0.082 |
|  | Global | -24.3±2.3 | -19.5±7.3 | 0.25 | -1.35±0.1 | -1.19±0.5 | 0.753 | 1.34±0.2 | 1.02±0.3 | 0.088 | 1.03±0.2 | 0.69±0.1 | 0.018* |

Group 3: abnormal, slow progression (C; initial LVEF<55%, +2 years ΔLVEF%≤5); Group 4: abnormal, fast progression (D; initial LVEF<55%, +2 years ΔLVEF%>5); \*p<0.05, adjusted for multiple comparisons.

## References

1. Landfeldt E, Thompson R, Sejersen T, McMillan HJ, Kirschner J and Lochmüller H. Life expectancy at birth in Duchenne muscular dystrophy: a systematic review and meta-analysis. European Journal of Epidemiology. 2020;35:643–653.

2. Crisafulli S, Sultana J, Fontana A, Salvo F, Messina S and Trifirò G. Global epidemiology of Duchenne muscular dystrophy: an updated systematic review and meta-analysis. Orphanet Journal of Rare Diseases. 2020;15.

3. Ryder S, Leadley RM, Armstrong N, Westwood M, De Kock S, Butt T, Jain M and Kleijnen J. The burden, epidemiology, costs and treatment for Duchenne muscular dystrophy: an evidence review. Orphanet Journal of Rare Diseases. 2017;12.

4. Meyers TA and Townsend D. Cardiac Pathophysiology and the Future of Cardiac Therapies in Duchenne Muscular Dystrophy. International Journal of Molecular Sciences. 2019;20:4098.

5. Shih JA, Folch A and Wong BL. Duchenne Muscular Dystrophy: the Heart of the Matter. Current Heart Failure Reports. 2020;17:57–66.

6. Angulski ABB, Hosny N, Cohen H, Martin AA, Hahn D, Bauer J and Metzger JM. Duchenne muscular dystrophy: disease mechanism and therapeutic strategies. Frontiers in Physiology. 2023;14.

7. Lee S, Lee M and Hor KN. The role of imaging in characterizing the cardiac natural history of Duchenne muscular dystrophy. Pediatric Pulmonology. 2021;56:766–781.

8. Earl CC, Soslow JH, Markham LW and Goergen CJ. Myocardial strain imaging in Duchenne muscular dystrophy. Frontiers in Cardiovascular Medicine. 2022;9.

9. Blaszczyk E, Gröschel J and Schulz-Menger J. Role of CMR Imaging in Diagnostics and Evaluation of Cardiac Involvement in Muscle Dystrophies. Current Heart Failure Reports. 2021;18:211–224.

10. Prakash N, Suthar R, Sihag BK, Debi U, Kumar RM and Sankhyan N. Cardiac MRI and echocardiography for early diagnosis of cardiomyopathy among boys with Duchenne muscular dystrophy: a cross-sectional study. Frontiers in Pediatrics. 2022;10:818608.

11. Lechner A, Herzig JJ, Kientsch JG, Kohler M, Bloch KE, Ulrich S and Schwarz EI. Cardiomyopathy as cause of death in Duchenne muscular dystrophy: a longitudinal observational study. ERJ Open Research. 2023;9:00176–2023.

12. Puchalski MD, Williams RV, Askovich B, Sower CT, Hor KH, Su JT, Pack N, Dibella E and Gottliebson WM. Late gadolinium enhancement: precursor to cardiomyopathy in Duchenne muscular dystrophy? The International Journal of Cardiovascular Imaging. 2009;25:57–63.

13. Earl CC, Pyle VI, Clark SQ, Annamalai K, Torres PA, Quintero A, Damen FW, Hor KN, Markham LW, Soslow JH and Goergen CJ. Localized strain characterization of cardiomyopathy in Duchenne muscular dystrophy using novel 4D kinematic analysis of cine cardiovascular magnetic resonance. Journal of Cardiovascular Magnetic Resonance. 2023;25.

14. Heidenreich PA, Bozkurt B, Aguilar D, Allen LA, Byun JJ, Colvin MM, Deswal A, Drazner MH, Dunlay SM, Evers LR, Fang JC, Fedson SE, Fonarow GC, Hayek SS, Hernandez AF, Khazanie P, Kittleson MM, Lee CS, Link MS, Milano CA, Nnacheta LC, Sandhu AT, Stevenson LW, Vardeny O, Vest AR and Yancy CW. 2022 AHA/ACC/HFSA Guideline for the Management of Heart Failure: A Report of the American College of Cardiology/American Heart Association Joint Committee on Clinical Practice Guidelines. Circulation. 2022;145.

15. Raucci FJ, Xu M, George-Durrett K, Crum K, Slaughter JC, Parra DA, Markham LW and Soslow JH. Non-contrast cardiovascular magnetic resonance detection of myocardial fibrosis in Duchenne muscular dystrophy. Journal of Cardiovascular Magnetic Resonance. 2021;23.

16. Cerqueira MD, Weissman NJ, Dilsizian V, Jacobs AK, Kaul S, Laskey WK, Pennell DJ, Rumberger JA, Ryan T and Verani MS. Standardized Myocardial Segmentation and Nomenclature for Tomographic Imaging of the Heart. Circulation. 2002;105:539–542.

17. Dann MM, Clark SQ, Trzaskalski NA, Earl CC, Schepers LE, Pulente SM, Lennord EN, Annamalai K, Gruber JM and Cox A. Quantification of Murine Myocardial Infarct Size using 2D and 4D High Frequency Ultrasound. American Journal of Physiology-Heart and Circulatory Physiology. 2022.

18. Damen FW, Salvas JP, Pereyra AS, Ellis JM and Goergen CJ. Improving characterization of hypertrophy-induced murine cardiac dysfunction using four-dimensional ultrasound derived strain mapping. American Journal of Physiology-Heart and Circulatory Physiology. 2021.

19. Leyba K, Paiyabhroma N, Salvas JP, Damen FW, Janvier A, Zub E, Bernis C, Rouland R, Dubois CJ, Badaut J, Richard S, Marchi N, Goergen CJ and Sicard P. Neurovascular hypoxia after mild traumatic brain injury in juvenile mice correlates with heart–brain dysfunctions in adulthood. Acta Physiologica. 2023;238.

20. Hor KN, Kissoon N, Mazur W, Gupta R, Ittenbach RF, Al-Khalidi HR, Cripe LH, Raman SV, Puchalski MD, Gottliebson WM and Benson DW. Regional Circumferential Strain is a Biomarker for Disease Severity in Duchenne Muscular Dystrophy Heart Disease: A Cross-Sectional Study. Pediatric Cardiology. 2015;36:111–119.

21. Kerstens TP, van Everdingen WM, Habets J, van Dijk AP, Helbing WA, Thijssen DH and Ten Cate FEU. Left ventricular deformation and myocardial fibrosis in pediatric patients with Duchenne muscular dystrophy. International Journal of Cardiology. 2023;388:131162.

22. Skaarup KG, Lassen MCH, Marott JL, Biering-Sørensen SR, Jørgensen PG, Appleyard M, Berning J, Høst N, Jensen G, Schnohr P, Søgaard P, Gislason G, Møgelvang R and Biering-Sørensen T. The impact of cardiovascular risk factors on global longitudinal strain over a decade in the general population: the copenhagen city heart study. The International Journal of Cardiovascular Imaging. 2020;36:1907–1916.

23. Grossman W, Jones D and McLaurin LP. Wall stress and patterns of hypertrophy in the human left ventricle. Journal of Clinical Investigation. 1975;56:56–64.

24. Siddiqui S, Alsaied T, Henson SE, Gandhi J, Patel P, Khoury P, Villa C, Ryan TD, Wittekind SG, Lang SM and Taylor MD. Left Ventricular Magnetic Resonance Imaging Strain Predicts the Onset of Duchenne Muscular Dystrophy–Associated Cardiomyopathy. Circulation: Cardiovascular Imaging. 2020;13.

25. Markham LW, Michelfelder EC, Border WL, Khoury PR, Spicer RL, Wong BL, Benson DW and Cripe LH. Abnormalities of Diastolic Function Precede Dilated Cardiomyopathy Associated with Duchenne Muscular Dystrophy. Journal of the American Society of Echocardiography. 2006;19:865–871.

